# Sensitive extraction-free SARS-CoV-2 RNA virus detection using a novel RNA preparation method

**DOI:** 10.1101/2021.01.29.21250790

**Authors:** Bin Guan, Karen M. Frank, José O. Maldonado, Margaret Beach, Eileen Pelayo, Blake M. Warner, Robert B. Hufnagel

**Author notes:** Corresponding author: Robert B. Hufnagel, Ophthalmic Genomics Laboratory, National Eye Institute, National Institutes of Health, 10 Center Drive, Building 10, Rm 10N109, Bethesda, MD, 20892, USA. Grant numbers and sources of support: Intramural Research Programs of the National Institutes of Health (Clinical Center, NEI, NIDCR).

## Abstract

Current conventional detection of SARS-CoV-2 involves collection of a patient sample with a nasopharyngeal swab, storage of the swab during transport in a viral transport medium, extraction of RNA, and quantitative reverse transcription PCR (RT-qPCR). We developed a simplified and novel preparation method using a Chelex resin that obviates RNA extraction during viral testing. Direct detection RT-qPCR and digital-droplet PCR was compared to the current conventional method with RNA extraction for simulated samples and patient specimens. The heat-treatment in the presence of Chelex markedly improved detection sensitivity as compared to heat alone, and lack of RNA extraction shortens the overall diagnostic workflow. Furthermore, the initial sample heating step inactivates SARS-CoV-2 infectivity, thus improving workflow safety. This fast RNA preparation and detection method is versatile for a variety of samples, safe for testing personnel, and suitable for standard clinical collection and testing on high throughput platforms.

## Introduction

Diagnostic testing for SARS-CoV-2 is crucial to combatting the current COVID-19 pandemic, even as vaccines are being distributed. The Centers for Disease Control and Prevention (CDC) protocol for detecting SARS-CoV-2 (COVID-19) involves sample collection, RNA extraction, and one-step real-time reverse transcription PCR (rRT-PCR or RT-qPCR) (accessed on 6/15/2020: https://www.fda.gov/media/134922/download). Uncontrolled community spread of SARS-CoV-2 has caused supply shortages for diagnostic tests, including the RNA extraction kits. Supply chain uncertainty and lack of adequate supplies remain widespread which hinders day-to-day laboratory operations and impedes efforts to increase testing capacity (survey results published on Nov. 9, 2020 and accessed on 11/12/2020: https://asm.org/Articles/2020/September/Clinical-Microbiology-Supply-Shortage-Collecti-1).

Recent research aimed at both simplifying current standard testing procedures for SARS-CoV-2 and alleviating RNA extraction kit shortage have shown encouraging results. First, specimens collected on nasopharyngeal swabs (NP swab) have been eluted into low-ethylenediaminetetraacetic acid (EDTA) tris buffer (lowTE) and directly used for one-step RT-qPCR without RNA extraction. This results in lower threshold cycles (Ct values).^1^ The lower Ct value suggests that eliminating the RNA extraction step may increase sensitivity. Secondly, samples collected in viral transport medium (VTM) have been successfully used for direct RT-qPCR detection after heating.^2-10^ Despite these noteworthy results, the Ct values were 4 or more cycles higher than the standard protocol utilizing the RNA extraction step. A lower inherent detection sensitivity leads to higher false negative rate, which is a particular risk for low viral load samples.^2, 5, 10^ Thirdly, proteinase K and heat has been used for NP and oropharyngeal swabs with 91% sensitivity, albeit with higher Ct than the conventional RNA extraction method in the RT-qPCR assays.^11, 12^ Recently, saliva samples with RNA extraction have been found to be an effective alternative sample type to NP swabs.^12-14^ Further studies showed that the SalivaDirect® method, employing proteinase K and heat-treatment, were able to detect 6 genome-copies per µl of SARS-CoV-2 without RNA extraction.^15^ While RNA extraction-free methods are promising, they lack the sensitivity and limit of detection compared to the current CDC protocol.

In this study, we aimed to improve the reported RNA extraction-free protocols to match or exceed the sensitivity to that of the standard protocol with RNA-extraction using both synthetic and human samples. We tested a variety of common chemicals that are used in molecular biology laboratories, and tested different preparation methods to identify reagents and conditions that (i) minimize RNA degradation, (ii) allow room temperature transport, heat inactivation, and storage of human samples, and (iii) provide sensitive RNA detection by RT-qPCR or RT-ddPCR. We found that Chelex^®^ 100, a chelating ion exchange resin, preserves essentially all SARS-CoV-2 RNA in the sample and permits direct detection by both RT-qPCR and RT-ddPCR. We further demonstrated a COVID-19 testing workflow that obliviate the use of a BSL-2 hood and omitting RNA-extraction while preserving the sensitivity of the standard protocol with RNA-extraction.

## Results

### Heating in lowTE buffer along with Chelex detects all virion RNA added

Given the promising results for COVID-19 testing obtained by direct elution of dried patient swab to lowTE (1x low-EDTA TE, 10 mM Tris, 0.1mM EDTA, pH8.0) buffer,^1^ and because heating has been shown to increase testing sensitivity for SARS-CoV-2 when samples stored in viral transport media (VTM) were used for direct testing without RNA extraction,^2, 5, 10^ we set out to test these conditions in a direct dried swab elution procedure. We simulated the dry swab procedure by desiccating the mixture of inactivated SARS-CoV-2 virions with known genome copies and 293FT cells using a speedvac centrifuge at room temperature. After resuspending the virus and cells in lowTE to a concentration of 1,000 genome copies/µl, we quantified the virus with RT-ddPCR using the N1 and N2 primers and showed that only ∼30% of virions were detected (Figure 1A, dried swab, no heat). Heating increased the detected virus to ∼40% of added virions (Figure 1A, dried swab, heat).

**Figure 1.**
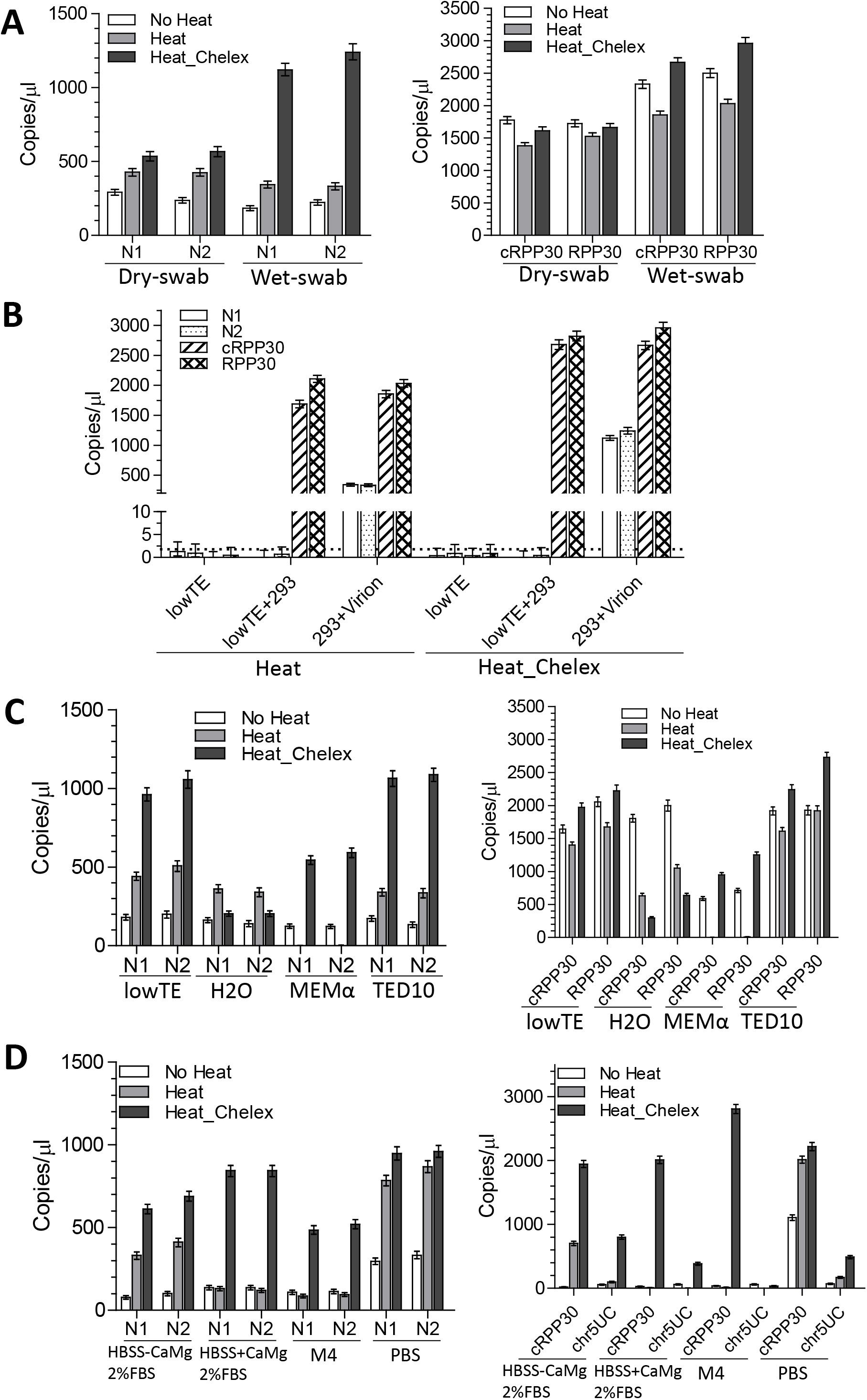
RT-ddPCR assays for simulated dry- and wet-swab using RNA-extraction free methods for SARS-CoV-2 detection. **(A)** 200,000 SASRS-Cov-2 virions and 20,000 293FT cells were dried at room temperature in a speedvac and then resuspend in 200 µl lowTE buffer to mimic the dry swab. The same amount of virions and 293FT cells directly added to 200 µl lowTE buffer was used to mimic the wet swab. The samples expected to have 1000 virion genome copies/µl were then used directly for RT-ddPCR (No Heat), heated at 98 °C for 5 min (Heat), or heated with 5% Chelex (Heat_Chelex). The RT-ddPCR reactions were carried out in one well for N1 and cRPP30 and another well for N2 and RPP30. **(B)** Negative controls and virion samples prepared as wet swab. The mean genome copies/µl of N1 & N2 were less than 1.2 in negative controls without virions added. N1 & N2 target SARS-CoV-2. cRPP30 is specific for *RPP30* cDNA, and RPP30 targets both genomic DNA and cDNA. Copies/µl refers to concentration in the samples used for RT-ddPCR. The error bars represent Poisson 95% confidence intervals. Dashed line indicates the threshold for the low detection limit of 1.8 copies/µl of SARS-CoV-2 virions. **(C)** Virions of 1000 genome copies/µl and 100 293FT cells/µl were prepared in lowTE, H2O, MEM alpha, or TED10, treated and assayed by RT-ddPCR as in (A). **(D)** Virions of 1000 genome copies/µl and 100 293FT cells/µl were prepared in HBSS with or without Ca^2+^ & Mg^2+^ supplemented with 2% FBS, M4, or PBS, treated and assayed by RT-ddPCR in for N1, N2, cRPP30 and chr5UC, a genomic DNA region on chromosome 5.

As RNases are Mg^2+^-dependent similar to DNases, and Chelex polymers have been used successfully for DNA preparation for its property of chelating divalent ions,^16^ we tested viral RNA isolation using Chelex. Heating the sample along with 5% Chelex improved virion detection to ∼50% (Figure 1A, dried swab, Heat_Chelex). We suspected that the desiccation-resuspension process caused viral and cellular RNA degradation. To confirm this, we added virions and 293FT cells directly to lowTE and measured viral RNA content (Figure 1A, wet swab). Heating the sample in lowTE in the presence of Chelex led to the detection of ∼100% of virions added. The *RPP30* mRNA also had the highest yield in the presence of Chelex. Heating without Chelex led to more *RPP30* mRNA degradation but better detection of the SARS-CoV-2 RNA as compared to no heat condition. The N1, N2 primers produced minimum background as shown in the controls (lowTE and 293FT cells suspended in lowTE) (Figure 1B).

The pH of 5% Chelex in water is 10-11.^16^ Because alkaline conditions are expected to lead to rapid RNA hydrolysis, we postulated that Tris pH8.0 in the lowTE buffer was critical for the high detection rate of SARS-CoV-2 virions in the presence of Chelex. To test this, we measured detectable virion RNA when samples were prepared in water (Figure 1C). In water, heating with Chelex only detected 20% of added virions, and 15% of the *RPP30* mRNA detected in the lowTE/Chelex condition. We also simulated swab stored in MEM α media, which has a similar formulation to VTM. Heating in MEM α led to an undetectable level of viral RNA and *RPP30* mRNA, possibly due to the 1.8 mM Ca^2+^ and 0.8 mM Mg^2+^ present in the media. Inclusion of Chelex during heating resulted in detection of >55% of viral RNA (Figure 1C). We found that inclusion of 2.5-5% DMSO in the final ddPCR reaction reduced negative droplets intensity (Supplementary Figure 1), we also simulated swab stored in the TED10 (90% lowTE + 10% DMSO) buffer, which results in 2.5% DMSO in the ddPCR reaction and thus potentially simplifies reaction setup. The viral and *RPP30* RNA amounts detected in the TED10 buffer in Chelex was slightly higher than those in lowTE buffer.

The CDC recommends VTM for specimens for viral culture and viral detection, which contains HBSS 1X with calcium and magnesium, 2% heat-inactivated FBS, Gentamicin and Amphotericin B (CDC SOP# DSR-052-05 accessed on 11/16/2020, https://www.cdc.gov/coronavirus/2019-ncov/downloads/Viral-Transport-Medium.pdf). The 1X HBSS buffer contains 1.3 mM Ca^2+^ and 0.9 mM Mg^2+^, which may lead to viral RNA degradation during heating, similar to MEM α medium. We tested the HBSS buffer without Ca^2+^/Mg^2+^ but supplemented with 2% heat-inactivated FBS, HBSS buffer containing Ca^2+^/Mg^2+^ and 2% heat-inactivated FBS, the M4 transport medium (HBSS based medium, containing HEPES, BSA, gelatin, and antibiotics), and PBS in the RNA-extraction free assay (Figure 1D). Heat increased viral detection for calcium/magnesium free media, but not the calcium/magnesium media. Only ∼12% of viral RNA was detected in the HBSS containing Ca^2+^/Mg^2+^ and 2% heat-inactivated FBS after heating, in contrast, heating in the presence of Chelex allowed the detection of 84% of virions.

In summary, LowTE pH 8.0 and TED10 with Chelex produced the highest amounts of viral RNA detected as compared to no heat or heating conditions among the buffers tested.

### Buffers allowing successful RT-qPCR without conventional RNA extraction

To investigate whether other agents besides Chelex could similarly be used for efficient RNA extraction-free SARS-CoV-2 detection by protecting against degradation, we tested various buffers for RT-qPCR without conventional RNA extraction. These buffers included RNA*later* (used for sample storage before RNA extraction), buffer RLT (lysis buffer from the Qiagen RNeasy RNA extraction kit), Urea (used for RNA extraction^17^), DMSO (used as lysis buffer for DNA extraction^18^ and known to inhibit RNases^19^), 1xTE (10 mM Tris, 1 mM EDTA, pH8.0), 10xTE (20mM Tris, 10 mM EDTA, pH7.5), and MEM α medium. Inactivated SARS-CoV-2 and 293FT cells were mixed and added to these buffers above along with lowTE and heated with or without Chelex (Figure S2A). We diluted the samples in water before RT-qPCR to avoid suspected inhibition of reverse transcription and PCR, and then compared the Ct values to calculated Ct values based on Ct of extracted RNA and number of virions present in each sample.

Of the chemicals tested, RNA*later* and RLT appeared to be incompatible with the RNA-extraction-free method, as there was either no amplification or the Ct values were much higher than expected for all four targets (Figure S2A) after 20-fold dilution. Urea, DMSO, TE or MEM α showed minimum RT-qPCR inhibition after dilutions. Chelex appears critical to achieve better detection of viral RNA and cellular RNA in DMSO, 1xTE, lowTE, or MEM α, with the average Ct cycle difference of N1 & N2 between Chelex and heat alone as 2.2, 1.8, 1.4, and 13, respectively. This represents improvement of sensitivity by Chelex of 4.5, 3.6, 2.7, and >1000 fold for samples prepared in DMSO, 1xTE, lowTE and MEM α, respectively. In summary, viral RNA detection sensitivity in simulated nasopharyngeal swab samples was highest in the presence of Chelex.

Next, we tested saliva samples with known added virion concentrations. Inactivated SARS-CoV-2 virions were also added to saliva, which were then mixed 1:1 with various buffers as above, to test whether saliva samples can be subjected to direct RNA detection. Saliva samples without exogenous chemicals or in DMSO were superior for RT-qPCR detection among the tested conditions (Figure S2A). Chelex 5% did not improve Ct for saliva, while decreased 3.4 Ct cycles for saliva 1:1 mixed with DMSO (Figure S2A).

We then determined the maximum concentrations of tested chemicals that were tolerated in a RT-qPCR reaction (Figure S2B). Samples were heat-treated with Chelex and serial dilutions of 2-fold were used for RT-qPCR. The highest chemical concentrations that did not interfere with RT-qPCR if using 5 µl of undiluted sample in a 20 µl reaction were Urea 0.5 M, DMSO 50%, EDTA 0.5 mM (Figure S2B). The N1 & N2 Ct values for the undiluted sample in lowTE was the lowest, lower than RNA-extraction using the same amount of virions, likely reflecting RNA loss during RNA extraction.

Together these results indicate that, by obviating the RNA extraction step, the presence of Chelex in sample buffer increases RNA molecules available for RT-qPCR, as we observed in a variety of buffers simulating nasopharyngeal and saliva collection conditions. Collecting swabs in lowTE appear to provide the highest sensitivity under synthetic conditions.

### Limit of detection

We attempted to further refine the buffer for this RNA-extraction free method by adjusting DMSO concentration and combining TE with DMSO. The RT-qPCR data showed that lowTE with heat and Chelex showed the lowest Ct values for N1 and N2, and combination of TE with DMSO did not improve the Ct (Figure S3A). Among the saliva samples, the undiluted saliva condition had the lowest Ct for N1 and N2 (Figure S3A), with Ct values ∼0.5 cycles above the lowTE sample. Heated saliva samples with or without Chelex lowered Ct values by more than 2 cycles compared to non-heated samples. We further found that increasing Chelex levels in DMSO increased detectable viral and cellular RNA (Figure S3B).

To improve the limit of detection for RT-qPCR, we optimized the reaction conditions by (i) including 2.5% DMSO in the final reaction if a sample does not contain DMSO, (ii) reducing the reaction volume to 10 µl, (iii) using the NEB-Luna-program II for RT-qPCR, reducing denaturing time from 10 seconds to 5 seconds and annealing/extension time from 40 seconds to 20 seconds (Figure S4). Under this condition, the Ct for Non-template controls were either undetermined or above 38. When analyzing the 10-fold serial dilutions of SARS-CoV-2 virions using our new preparation method by RT-qPCR, the LoD was 200 copies per swab or 1 copy/µl of samples before RT-qPCR (Figure 2A). Using the conventional RNA extraction method, the LoD was 2,000 genome copies per swab (Figure 2A). The LoD for saliva samples using the Chelex method was also at 1 copy/µl for all replicates when 50 µl whole unstimulated saliva was added to 25 µl of 50% Chelex, compared to the proteinase K method, which was undetermined for a single 1 copy/µl replicate (Figure 2B).

**Figure 2.**
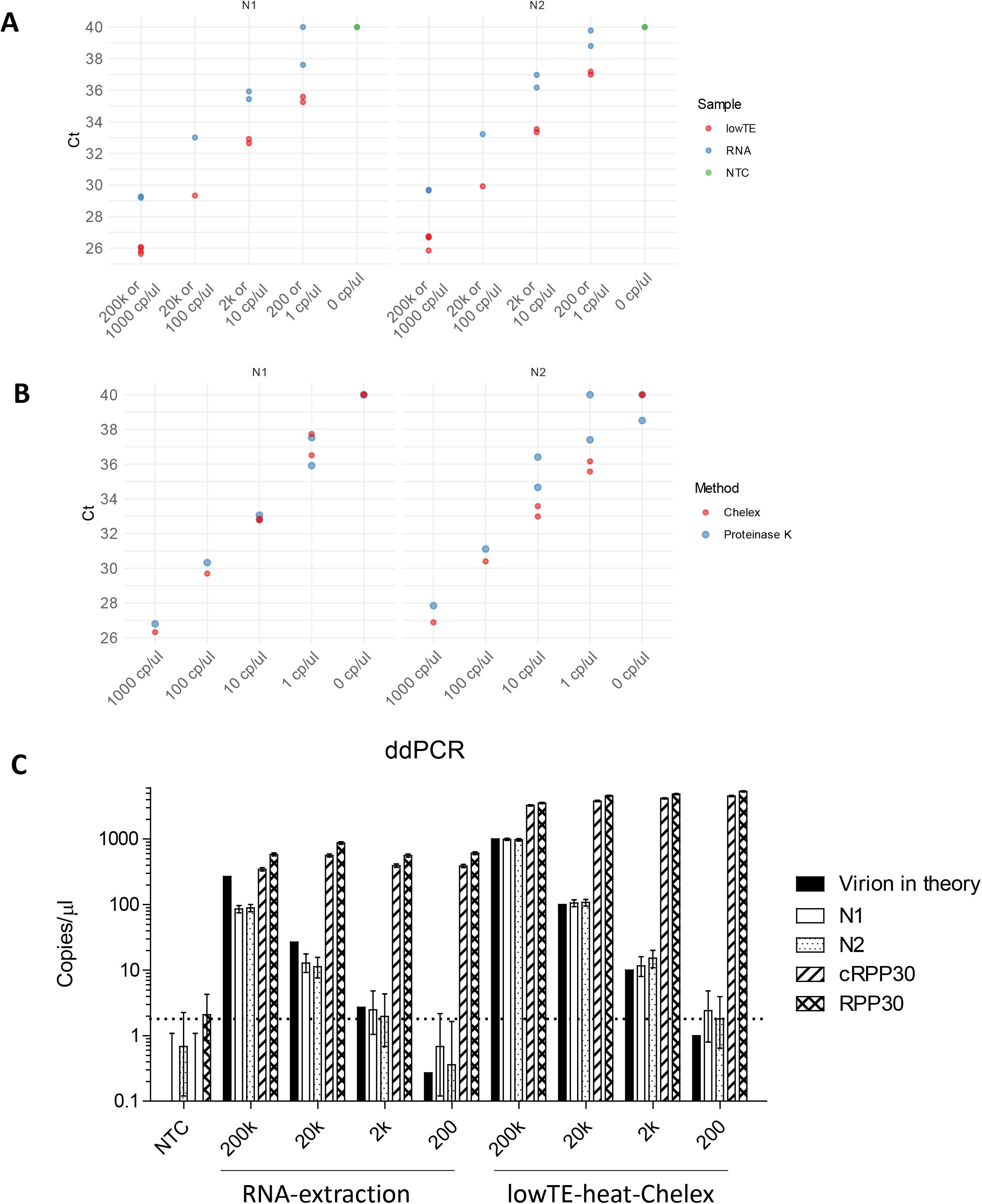
The limit of detection of SARS-CoV-2 using RT-qPCR or -ddPCR. **(A)** RT-qPCR comparing Chelex-RNA and conventional RNA extraction. RNA refers to RNA prepared by simulating conventional method with RNA-extraction: a swab with 200,000 to 200 genome copies of SARS-CoV-2 virions was added to 3 ml of VTM, of which 200 µl were used for RNA extraction, and RNA was eluted in 50 µl H2O. LowTE refers to simulating a swab with 200,000 to 200 genome copies of virions eluted in 200 µl lowTE, and then heated in the presence of 5% Chelex. **(B)** RT-qPCR comparing Chelex and Proteinase K methods for saliva samples. 1000 to 1 genome copies/µl of SARS-CoV-2 virions were prepared in saliva samples and subjected to the Chelex or Proteinase K methods and RT-qPCR. The NEB Luna RT-qPCR kit and NEB-Luna-Program II was used with 2.5 µl samples in 10 µl reaction volume. Samples with undetermined Ct values were plotted as Ct 40. **(C)** The limit of detection of SARS-CoV-2 using RT-ddPCR. Samples from (A) were used for RT-ddPCR. 5 µl of sample was used for RT-ddPCR in 20 µl reaction volume. The mean genome copies/µl of N1 & N2 were less than 1.2 in negative controls without virions added. N1 & N2 target SARS-CoV-2. cRPP30 is specific for *RPP30* cDNA, and RPP30 targets both genomic DNA and cDNA. Copies/µl refers to concentration in the samples used for RT-ddPCR. The error bars represent Poisson 95% confidence intervals. Dashed line indicates the threshold for the low detection limit of 1.8 copies/µl of SARS-CoV-2 virions. NTC, no-template control.

We then used RT-ddPCR to determine whether we could detect a lower virion copy number. Because we occasionally observed 1 or 2 positive droplets in the no-template control reactions, we applied 1.8 copies/µl, or 50% higher than the maximum N1/N2 mean of negative controls (Figure 1B and Figure 2C), for the mean of N1 and N2 as the threshold for being positive for the SARS-CoV-2 N1/N2 RT-ddPCR assay. RT-ddPCR confirmed the LoD for conventional RNA extraction method to 2,000 virions per swab and the lowTE-Heat-Chelex method to 200 virions per swab (Figure 2C and Table 1). The RNA extraction method only detected 30-50% of RNA molecules at the viral loads tested (Table 1).

**Table 1.** Limit of Detection comparing conventional RNA extraction and new preparation method

| “Swab”<br>viral load<br>(genome<br>copies) | RNA-extraction |  |  | lowTE_Heat_Chelex |  |  |
| --- | --- | --- | --- | --- | --- | --- |
|  | Theoretical<br>virus<br>concentration*<br>(copies/μl) | Mean of<br>N1/N2<br>concentration<br>(copies/μl) | Percentage<br>detected | Theoretical<br>virus<br>concentration*<br>(copies/μl) | Mean of<br>N1/N2<br>concentration<br>(copies/μl) | Percentage<br>detected |
| **200k** | 266.7 | 86.8 | 33% | 1000 | 976 | 98% |
| **20k** | 26.7 | 12.0 | 45% | 100 | 106 | 106% |
| **2k** | 2.7 | 2.2 | Detectable<sup>#</sup> | 10 | 13.4 | 134% |
| **200** | 0.3 | 0.5 | ND<sup>#</sup> | 1 | 2.1 | Detectable<sup>#</sup> |

\*: Virus concentration in samples used for RT-ddPCR if there is no loss during sample processing.

In summary, the lower limit of detection of the Chelex method is 1 copy/µl in NP or saliva under optimized buffer conditions and it avoids virion loss as observed in RNA extraction protocols.

### Patient samples

We then validated our Chelex RNA preparation method using primary patient samples. NP swabs were collected in M4 (N=14, S01 to S14, Figure 3A) or PBS (N=2, S15 & S16). These samples were tested in the NIH Clinical Center diagnostic laboratory using conventional CDC RNA extraction and RT-qPCR method (easyMAG-CL method), then frozen. Three of these samples, S01 to S03, had viral titer above 200 genome copies/µl. Twelve samples had viral titer less than 20 genome copies/µl, including eight considered indeterminate because only one of N1 or N2 targets was positive. One sample, S14, was negative.

**Figure 3.**
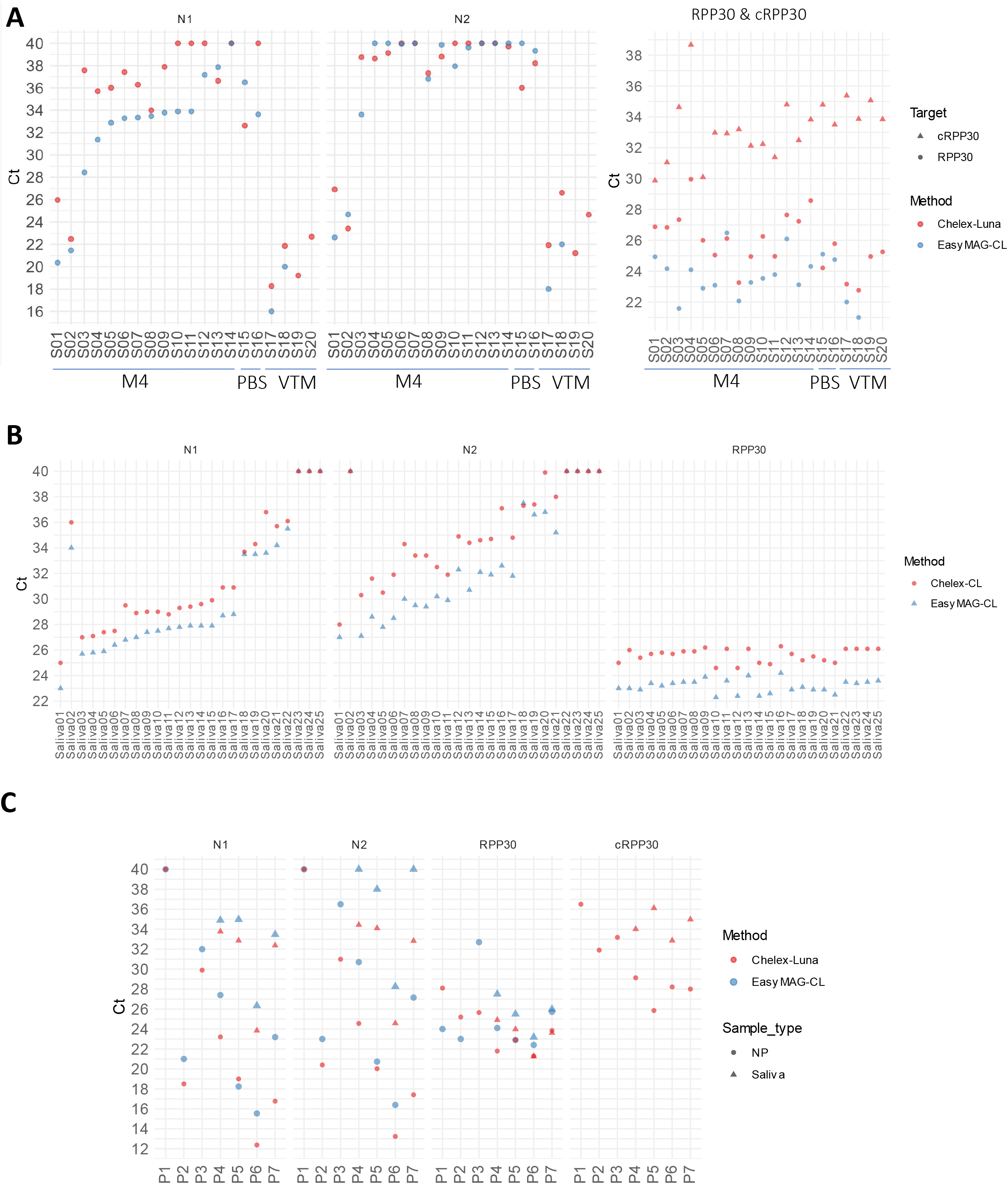
SARS-CoV-2 detection of patient samples prepared by the Chelex method. **(A)** Patient NP swab samples were heated in the presence of 5% Chelex (S01 to S16, and S19, S20) or 10% Chelex (S17 & S18). S19 & S20 are 1:2 dilution of S17 & S18 in LowTE, respectively. **(B)** 50 µl of patient saliva samples (Saliva01 & 02) or negative patient saliva samples spiked with positive patient saliva samples (Salia03 to 22) were mixed with 25 µl of 50% Chelex in TED99, and heated for 5 min in a ThermoMixer. **(C)** Paired NP swabs from seven patients (P1 to P7) and saliva-saturated swabs from four patients (P4 to P7) were collected in VTM or Chelex collection tubes. VTM samples were used for RNA extraction (EasyMag). Luna refers to the NEB Luna RT-qPCR kit and NEB-Luna-Program II with 2.5 µl samples in a 10 µl reaction volume. CL refers to CDC assay performed in the clinical laboratory with 5 µl samples in a 20 µl reaction volume. Undetermined Ct values were plotted as Ct 40.

Samples were thawed and mixed with the Chelex resin, heat-inactivated, and subjected to RT-qPCR using the NEB Luna RT-qPCR kit (Chelex-Luna method). As the easyMAG-CL method enriched sample 4-fold before RT-qPCR, we expected that Ct values from the Chelex-Luna method would increase by 2-fold including loss that may occur during RNA extraction (50% detectable viral materials in M4 in the Chelex-Luna assay; Figure 1D). The Ct values for N1, N2, and RPP30 from clinical laboratory RT-qPCR and NEB Luna RT-qPCR were comparable when using a common set of purified patient RNA samples with different viral titer (data not shown). The mean difference of N1 Ct between the two methods was 2.7, excluding four samples (S10, S11, S12, and S16) that did not show a Ct value in the Chelex-Luna assay.

Of the twelve samples with less than 20 genome copies/µl, eight (or 67%) showed as positive in the N1 Chelex-Luna assay, including two samples (S13 and S15) that showed lower Ct values and another (S08) with the similar Ct value. The N2 Ct values were higher than N1 and many were higher than 38, thus were not informative for the low titer samples in the Chelex-Luna assay. Two NP swab samples stored in the CDC-suggested VTM (HBSS with Calcium & Magnesium, S17 and S18) were tested side-by-side using the easyMag-CL and Chelex-Luna methods. The N1 & N2 Ct values of both samples increased 2 and 4, respectively, reflecting likely inhibition by the VTM as observed in the synthetic samples (Figure 3A).

Next, we compared the RNA extraction and Chelex methods using two primary saliva samples (Saliva01 & 02) and 20 positive saliva samples diluted in negative saliva samples side-by-side (Saliva03 to 22, Figure 3B). The mean Ct differences between the Chelex and RNA extraction methods for N1 and N2 were 1.6 and 2.6 respectively. Among the six samples with less than 10 genome copies/µl as determined by the RNA extraction method, five showed as positive and one (Saliva20) was indeterminate in the Chelex assay (Figure 3B). Thus, the Chelex method demonstrated similar sensitivity as the RNA extraction method for both primary NP swab and saliva samples.

We then tested the optimized Chelex RNA isolation method prospectively by collecting symptomatic patient NP swabs (n = 7) and saliva-saturated swabs (n = 4) directly into 0.5 ml (NP) or 0.4 ml (saliva) Chelex containing buffer, side-by-side with swabs collected into 3 ml VTM. The Chelex samples were heated and used for NEB Luna RT-qPCR, and the samples in VTM were subjected to RNA extraction followed by RT-qPCR performed at the clinical laboratory. Because 1/6^th^ of buffer in the collection tube was used in the Chelex method and RNA extraction concentrated sample by 4-fold, the Ct values in the Chelex method are expected to be 0.6 lower than the RNA extraction method. Sample P1 was found to be negative using both methods, and five of the six NP samples and four of the four saliva samples had lower N1 and N2 Ct values in the Chelex method as compared to the RNA extraction method (Figure 3C). The NP sample N1 Ct differences for patients P2 to P7 between these two methods were −1.9, −2.1, −4.2, 0.8, −3.2, and −6.4, and their N2 Ct differences were −0.3, −5.5, −6.1, −0.7, −3.2, and −9.7. The saliva samples’ N1 Ct differences for patients P4 to P7 between these two methods were −1.1, −2.1, −2.5, and −1.1, and their N2 Ct differences were −5.6, −3.9, −3.7, and −7.2. Thus, the Chelex method may offer better sensitivities using the procedure here. In addition, the Chelex method allowed sample processing without a Biosafety Cabinet hood as the samples were inactivated before tube opening.

### Sample stability at room temperature

We then determined whether viral and cellular RNA were stable over time. Samples were stored at room temperature then heat-treated in the presence of Chelex and assayed at different timepoints as indicated (Figure 4). Viral RNAs in lowTE or TED10 were relatively stable at room temperature and ∼80% of N1 or N2 RNAs were detected on day 5 (Figure 4A). Viral RNAs were less stable in MEM α medium as > 80% were degraded after 3 days (Figure 4A). The cellular RNA was less stable than viral RNA in the similar conditions (Figure 4A). We then determined the RNA stability at room temperature after heat-treatment with Chelex (Figure 4B). Heat-treatment stabilized both viral and cellular RNA, and >80% viral RNAs and >60% cellular RNA were detected on day 5 (Figure 4B).

**Figure 4.**
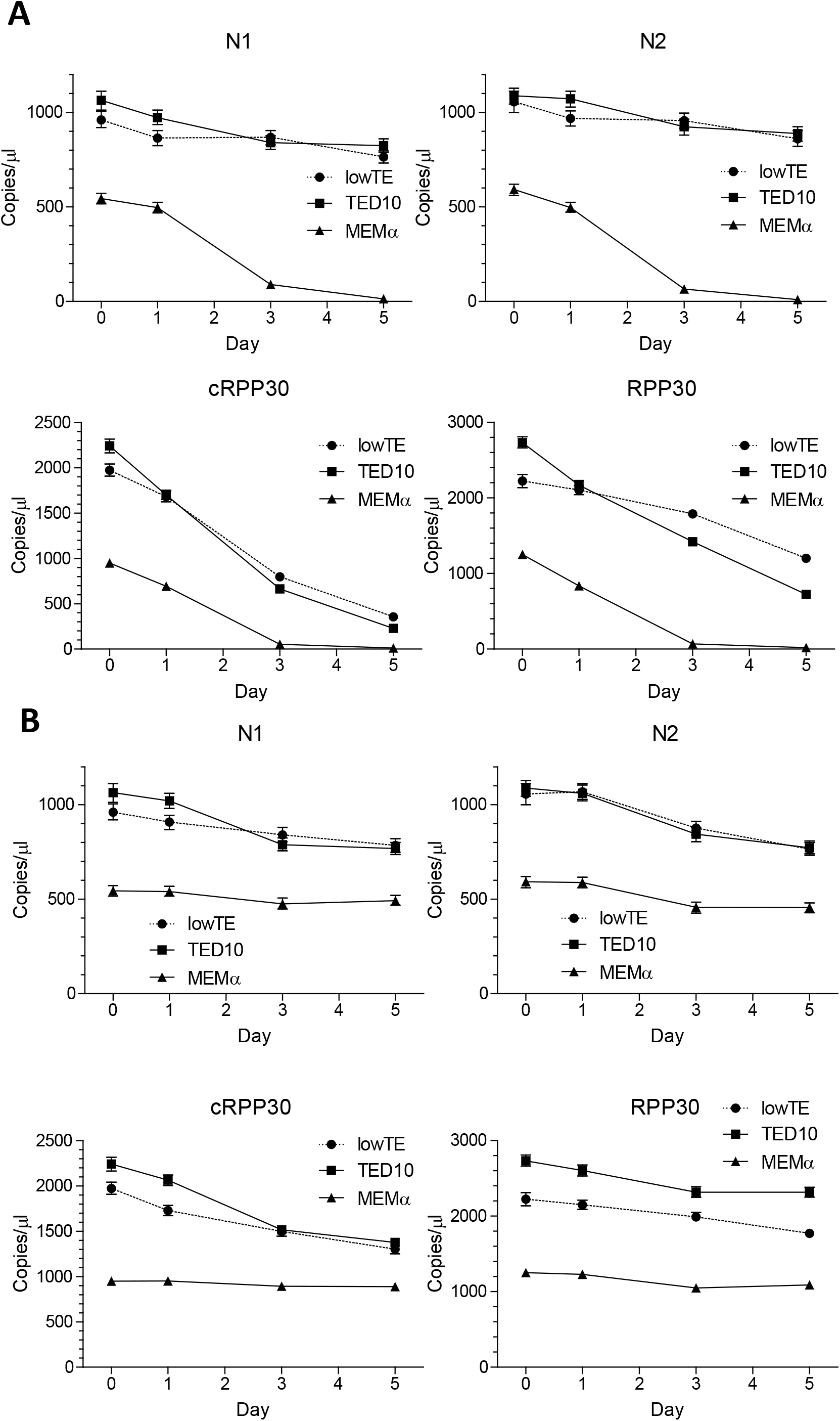
Viral and cellular RNA stability in lowTE by RT-ddPCR assays. **(A)** Virions of 1000 virion genome copies/µl and 100 cells/µl 293FT cells were prepared in lowTE, TED10 or MEM α, and stored at room temperature. Samples were heated with 5% Chelex on the time points indicated and assayed. The RT-ddPCR reactions were carried out in one well for N1 and cRPP30 and another well for N2 and RPP30. **(B)** Virions of 1000 virion genome copies/µl and 100 cells/µl 293FT cells were prepared in the buffers, heated with 5% Chelex on day 0, and assayed on a time series. Copies/µl refers to concentration in the samples used for RT-ddPCR. The error bars represent Poisson 95% confidence intervals.

A similar stability experiment was performed for virions prepared in saliva samples. The viral RNAs were stable in saliva before heat-treatment, as a higher amount of viral RNAs were detected after storage at room temperature, possibly because interfering agents in saliva degrade during storage (Figure 5A). However, the viral RNA stability decreased markedly after heat treatment (Figure 5B).

**Figure 5.**
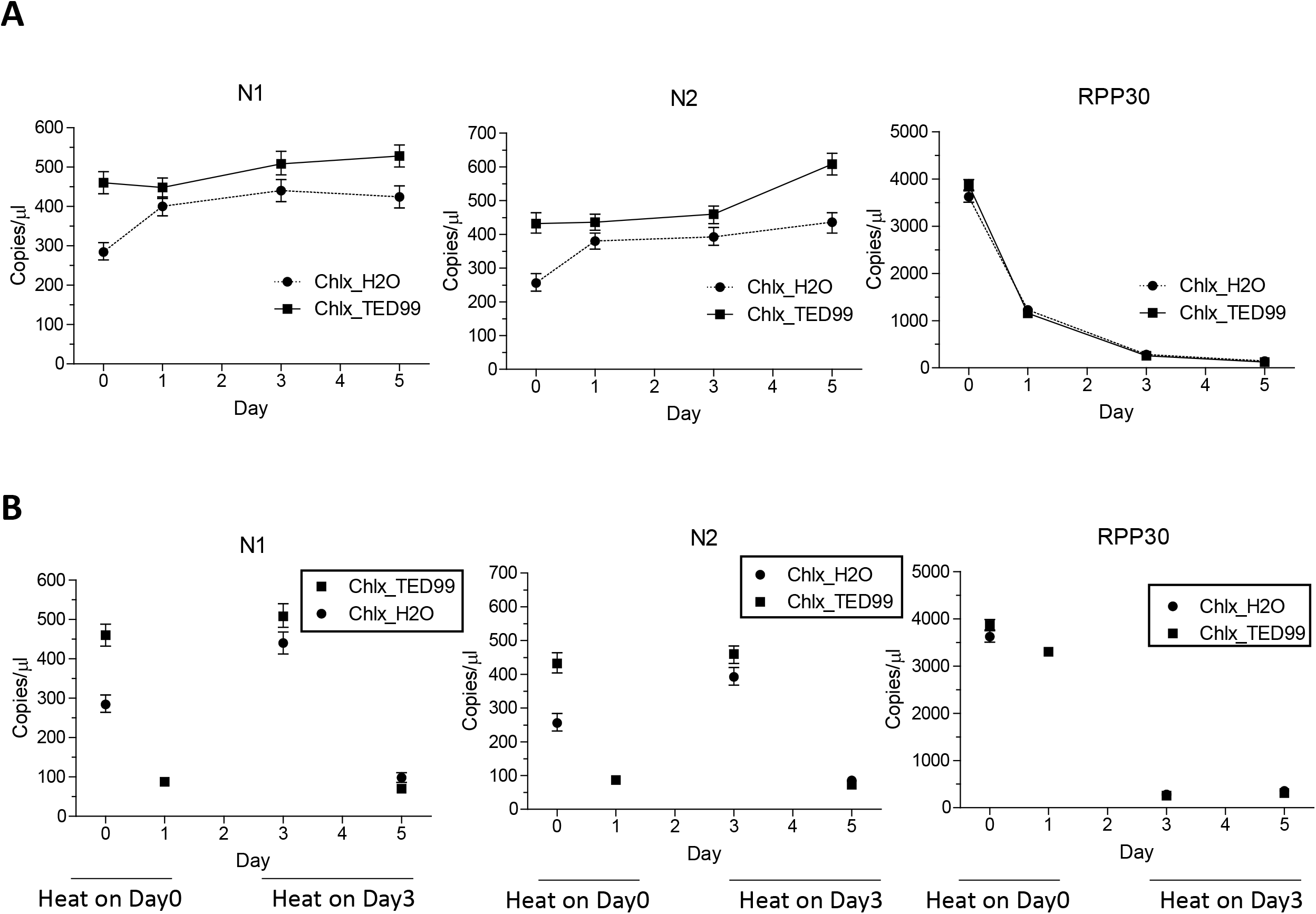
Viral RT stability in saliva by RT-ddPCR assays. **(A)** Virions were added to saliva samples at 1000 virion genome copies/µl and stored at room temperature. Samples were heated with 1/5 volumes of 50% Chelex prepared in H2O or TED99 on the time points indicated and assayed. The RT-ddPCR reactions were carried out in one well for N1 and cRPP30 and another well for N2 and RPP30. **(B)** Virions of 1000 virion genome copies/µl were prepared in saliva as in (A), heated with Chelex on day 0 or day 3, and assayed on the days indicated. cRPP30 data points were not plotted because of the low level detected. Copies/µl refers to concentration in the samples used for RT-ddPCR. The error bars represent Poisson 95% confidence intervals.

## Discussion

Our novel preparation approach using Chelex polymer increased RNA yield available for RT-qPCR and RT-ddPCR detection compared to the conventional method using the simulated samples with the ATCC SARS-CoV-2 virions. When using stored patient samples, RNA extraction provided higher sensitivity (Figure 3A, B). This is likely due to the enrichment arising from the lower volume of RNA eluate as compared to the input sample volume. The lower sensitivity of the Chelex method seen in Figure 3A could be also due to RNA loss during the freeze-thaw cycle. When the samples were tested side-by-side (Figure 3B), the sensitivities of the Chelex method and RNA extraction method are comparable. When patient samples were collected in VTM (S17 & S18 in Figure 3A), the N1 and N2 Ct values increased 2 and 4, respectively, suggesting that the Chelex method on VTM-collected samples may result in lower sensitivity for some low viral load samples. Further, we found that N1 and N2 responded differently toward components in the VTM as the N1 Ct increased while N2 Ct decreased after 1:2 dilution (S19 & S20 in Figure 3A). Our results in Figure 3C showed that collecting the swab directly in Chelex-TED (TE+DMSO) buffers followed by heating provided the best sensitivity for SARS-CoV-2 detection. The higher LoD associated with conventional RNA extraction method was due to dilution in the VTM and loss of RNA during RNA extraction as shown in Table 1. In addition, viral RNA may be degraded faster when storing in the calcium/magnesium containing VTM, as suggested by data presented in Figure 4A. As such, we recommend primary sample collection for this RNA isolation method in either TED or lowTE buffer, which has similar or better detection performance as RNA extraction methods for primary samples collected in VTM.

We also demonstrate that the Chelex method allows for highly-reproducible detection of 1 genome copy/µl of the SARS-CoV-2 virions, a 6-15-fold improvement in detection sensitivity.^15, 20^ One explanation for this improvement is that the Chelex resin chelates the divalent ions necessary for Rnase activities, and the resin may be able to non-specifically remove inhibitors to reverse-transcription and/or PCR. In addition, un-processed samples may lead to more RNA degradation during sample collection and storage as compared to being stored in the presence of Chelex. We further identified conditions that allow sensitive detection of SARS-CoV-2 in saliva. The potential benefits of saliva testing include lower cost (no swab), reduced variability, and improved patient acceptance over traditional NP swab.^14, 21^ Thus, the Chelex method may provide a more sensitive point-of-care method for RNA diagnostics by reducing false negative results.

There will be a continued need for SARS-CoV-2 detection during and after the current vaccination period. The sheer volume of preprint and publications in a short period of time illustrates the urgent need and hope to increase testing capacity employing the RNA-extraction free approach. The utility of this RNA isolation method for both NP and saliva samples would increase the number of people tested in the same timeframe as the current method. In addition to improved sensitivity, this method offers a number of additional advantages compared to the current gold standard clinical laboratory testing, including improved cost, reduced sample processing time and complexity, and enhanced workflow safety. The cost of CDC-recommended VTM collection tube is ∼$1.70 per tube, and RNA extraction may cost > $6 per sample. We estimate the total cost of Chelex, lowTE, DMSO, and the collection tube is <$1 per sample. Thus, the Chelex method will save cost and reduce supply chain burden by eliminating the need for RNA extraction and VTM. The novel RNA preparation method, amenable for high-throughput processing, is expected to shorten diagnostic testing time by omitting the RNA extraction step and omitting the chemical disinfection of patient samples. This method utilizes a heat-inactivation step that minimizes viral RNA loss, obviates the need for a biological safety cabinet, and eliminates exposure of laboratory personnel to live virus. Therefore, we fully expect that this method will facilitate broader availability and testing capacity for not only, COVID-19, but also for other infectious pathogens. Because of the observed stability of SARS-CoV-2 RNA in collected samples at room temperature, this method should also improve access to COVID-19 testing in resource scarce regions of the world, by improving RNA stability, reducing cost of collection kits and diagnostic reagents, and eliminating the requirement of refrigeration, biosafety cabinet, and storage of RNA extraction kits.

Because the Chelex method also allowed cellular RNA detection, we expect that the method could find wide use in both clinical and research laboratories. DNA present in the solution is not expected to interfere with many applications because RT-qPCR can be performed using exon-spanning primers and polyA selection is often an integrated step in RNAseq. The DNA present may be also used as a normalization control since the DNA content reflects the cell number when two cell populations have similar percentages of cells in the G2/M phases. This helps alleviate the concerns of choosing a proper house-keeping gene during RNA expression analysis.^22^ If DNA needs to be removed before downstream application, Dnase I treatment may be performed using commercially available Dnase I kits. The new RNA preparation method could also be used together with other RNA detection method such as rolling circle amplification, loop-mediated isothermal amplification or SHERLOCK.^23-25^

One limitation of the current study was the low-level contamination observed in the RT-ddPCR assay, where one or two positive droplets were observed in the no-template control reactions. The contamination could result from either viral RNA or PCR amplicon present in the research laboratory. Due to the background contamination, we used 1.8 copies/µl as the low limit of detection for the SARS-CoV-2 N1/N2 RT-ddPCR assay. According to the FDA Emergency Use Authorization (EUA) by the manufacturer Bio-Rad, the low limit of detection in the 1-well RT-ddPCR assay system is 2 positive droplets and no positive droplets observed in no-template controls. Thus the low limit of detection in a RT-ddPCR assay could reach 0.1 copy/µl if performing in a clean room.

In summary, we robustly demonstrate improvements in COVID-19 viral testing workflow using synthetic and real-world samples employing the Chelex-based extraction-free workflow. This methodology has the clear benefit of dramatic improvements in sensitivity, cost, and time savings for clinical laboratory testing. Additionally, this method exhibits improved safety characteristics including obviating the need for the use of a biological safety cabinet and harsh disinfection chemicals prior to testing. Finally, this method is easily adapted to both clinical and research laboratories and could be standard of care for nucleic acid testing and transport in the near future.

## Online Methods

### Human samples

Samples were collected from healthy volunteers and subjects who provided informed consent to National Institutes of Health (NIH) Institutional Review Board (IRB)-approved protocols (20-D-0094, NCT04348240; 20-CC-0128, NCT04424446) at the NIH Clinical Center.

### Sample collection

We used four types of samples for assay development and validation: (i) simulated samples containing the heat-inactivated SARS-CoV-2 virions obtained from ATCC (VR-1986HK, concentration measured by ddPCR at ATCC); (ii) positive patient saliva samples diluted into negative saliva samples; (iii) historical patient samples including nasopharyngeal swab and whole unstimulated saliva; and (iv) prospective paired nasopharyngeal and saliva samples swabs. The paired NP and saliva samples were collected at the NIH Symptomatic Testing Facility. Briefly, a FLOQswab® (COPAN) was inserted into the nares and advanced posteriorly to the nasopharynx as previously described.^26^ Swabs were then inserted into VTM. Subjects were asked to expectorate into a sterile 50 mL conical vial every 30 seconds for 3-5 minutes, or until ∼3 mL of saliva was collected. A FLOQswab^®^ was inserted into the saliva and immediately transferred into VTM. Samples were placed on ice and shipped immediately to the NIH Clinical Center Clinical Laboratory for standard CDC testing. In parallel, NP samples were placed in 0.5 ml 10% Chelex in TED10 buffer (90% lowTE and 10% DMSO), and swabs with saliva were placed in 0.4 ml Chelex saliva buffer. The 0.4 ml Chelex saliva buffer was made by mixing 0.2 ml of 50% Chelex in TED99 and 0.2 ml of lowTE. TED99 was 10mM Tris pH8.0, 0.1mM EDTA and 99% DMSO, which was made by mixing 100 µl of 1 M Tris pH8.0, 2 µl of 0.5 M EDTA, and 9.9 ml of DMSO. These patient samples with Chelex in 15-ml tubes were heat-inactivated in a thermomixer for 10 min at 98 °C before testing.

### Clinical laboratory CDC SARS-CoV-2 Assay

The conventional RNA-extraction RT-qPCR method following CDC guideline has been reported, and herein referred as easyMAG-CL (clinical laboratory) assay.^21^ Briefly, nucleic acid from individual specimens was extracted from 200 μL of Saliva/NPspecimens using the NucliSENS^®^ easyMAG^®^ platform (bioMérieux, Marcy l’Etoile, France) with an elution volume of 50 μL. 5 µl RNA was then used for RT-qPCR in 20 µl reaction volume. PCR was performed on the ABI 7500 Fast Real-Time PCR System (Thermo Fisher Scientific, Waltham, MA). The assay utilized primer/probe sets for nucleocapsid protein, 2019-nCoV_N1 and 2019-nCoV_N2, and the human RNase P (*RPP30* gene, also referred as *RP* gene in the CDC protocol) as an internal control to ensure that extraction and amplification was adequate as described. Cycle threshold (Ct) values were recorded for N1, N2 and RNAse P for each sample. Samples were considered positive for SARS-CoV-2 when both N1 and N2 targets were detected with Ct count <40. The positive signal for N1 or N2 alone was defined as an indeterminate result.

### Chemicals, buffers, and cells

Chelex® 100 Resin (hereafter “Chelex”; cat# 142-1253, molecular biology grade) was obtained from Bio-Rad. Molecular biology grade water (cat# 351-029-101), TE pH 8.0 (cat# 351-011-131), 1M Tris-HCl pH7.5 (cat# 351-006-101), HBSS with Ca^2+/^Mg^2+^ (cat# 114-061-101) were from Quality Biological (Gaithersburg, MD). The low-EDTA TE or lowTE buffer (10 mM Tris, pH8.0, 0.1 mM EDTA, cat# 12090-015), Urea (cat# 15505-050), RNAlater (cat# AM7024), MEM αmedium (cat# 12571-063), HBSS without Ca^2+/^Mg^2+^ (cat# 14175-095), M4 (cat# R12550), heat-inactivated FBS, 293FT cells were procured from Thermo Fisher Scientific. Dimethyl sulfoxide (DMSO, cat# D2650-100ML) was from Sigma. 1M Tris-HCl pH8.0 (cat#221-232) and 0.5M EDTA (cat#221-057) were from Crystalgen (Commack, NY). PBS (cat# RGF-3190, pH7.2) was from KD Medical (Columbia, MD). TED10 buffer was 90% lowTE and 10% DMSO (volume to volume).

Chelex was first prepared in H2O, lowTE, or TED99 at 50% (50 grams/100 ml total volume, or 500 milligrams Chelex to 550 µl H2O). The 50% Chelex was then added in 1/10 volume to samples to obtain 5% Chelex in a PCR strip with a wide-bore tip. The samples were vortexed briefly then heated in a PCR cycler for 5 min at 98 °C, followed by spinning at 1,000 to 2,000xg for 2 mins in a swing-rotor. The supernatant was then used for RT-qPCR or -ddPCR.

For simulated samples involving the ATCC virions, the RNeasy mini kit (Qiagen) was used to extract RNA from virion and cell mixture of less than 10 µl that contained the expected amounts of virions. The RNeasy Protect Saliva Mini Kit (Qiagen) was used for RNA extraction from ATCC virion-simulated saliva samples.

### Primers, RT-qPCR and RT-ddPCR

The primer and probe sequences used for detecting SARS-CoV-2 and human *RPP30* targets in the RT-ddPCR and NEB Luna RT-qPCR were identical to the easyMAG-CL assay above. The *RPP30* (or RP) primers and probe in the CDC protocol, whose sequences are RPP30F AGATTTGGACCTGCGAGCG, RPP30R GAGCGGCTGTCTCCACAAGT, RPP30Hex TTCTGACCTGAAGGCTCTGCGCG, amplify the DNA sequence located in the exon 1 of the *RPP30* gene, thus are expected to amplify both cDNA and genomic DNA contents. An additional *RPP30* primer specific for *RPP30* cDNA was designed to span the exon 1 and exon 2, RPP30cR GCAACAACTGAATAGCCAagGT, where the lower case “ag” denotes the exon junction. RPP30cR was used for RT-qPCR or -ddPCR together with RPP30F and RPP30Hex. The primer and probe sequences for an ultra-conserved region in chromosome 5 are: chr5UC-F ATTTATGACCAGCCACAGCC, chr5UC-R CCATCAGGGACTTGGTTTCA, chr5UC-Hex CAACTCCAGCAGCTGCACACCGC. Primers and probes were ordered from Eurofins Genomics or IDTDNA.

The Luna Universal Probe One-Step RT-qPCR Kit (#E3006X, NEB) was used for RT-qPCR with the following cycling conditions using a QuantStudio 3 real-time PCR system (ThermoFisher Scientific): NEB-Luna-Program I: 55°C for 10 min, 95°C for 1 min, and 45 cycles of 95 °C for 10 sec and 60 °C for 40 sec or NEB-Luna-Program II: 55°C for 10 min, 95°C for 1 min, and 45 cycles of 95 °C for 5 sec and 60 °C for 20 sec. Ct (Crossing threshold) values for N1 & N2 (viral targets) were set at 0.1 ΔRn, Ct values for cRPP30 (specific for RPP30 cDNA) and RPP30 (targets both genomic DNA and cDNA) were at 0.02 ΔRn. The N1 and N2 Ct of less than 38 were considered positive in the NEB Luna RT-qPCR assay.

The 1-Step RT-ddPCR Advanced Kit for Probes (#186-4021, Bio-Rad) was used for RT-ddPCR using the QX200 Droplet Digital PCR System (Bio-Rad). The cycling condition for RT-ddPCR was: 50°C for 60 min, 95°C for 10 min, and 40 cycles of 94°C for 10 sec and 55°C for 60 sec, followed by 98°C for 10 min, 4°C for 30 min then hold at 4°C. If DMSO was not present in the sample, DMSO was added to 2.5% in the RT-qPCR or -ddPCR reaction, unless specified otherwise.

## Data Availability

Methodology paper, thus not applicable here.

## Acknowledgments

This work was supported by the Intramural Research Programs of the National Institutes of Health (Clinical Center, NEI, NIDCR). The content of this publication does not necessarily reflect the views or policies of the Department of Health and Human Services, nor does mention of trade names, commercial products, or organizations imply endorsement by the U.S. Government. We would like to acknowledge staff in the National Institutes of Health Department of Laboratory Medicine, Occupational Medical Services COVID-19 Testing Facility, and the Division of Fire and Rescue Services.

## Author contributions

B.G., K.M.F., B.M.W. and R.B.H. designed experiments and wrote the manuscript. K.M.F., J.O.M., M.B., E.P., and B.M.W. collected patient samples. All authors contributed to the manuscript editing.

## Competing interests

NEI (B.G. and R.B.H.) filed an invention disclosure. NEI has protected the intellectual property around this technology which is available for licensing and co-development. Please contact for more information.

---

**Figure S1.**
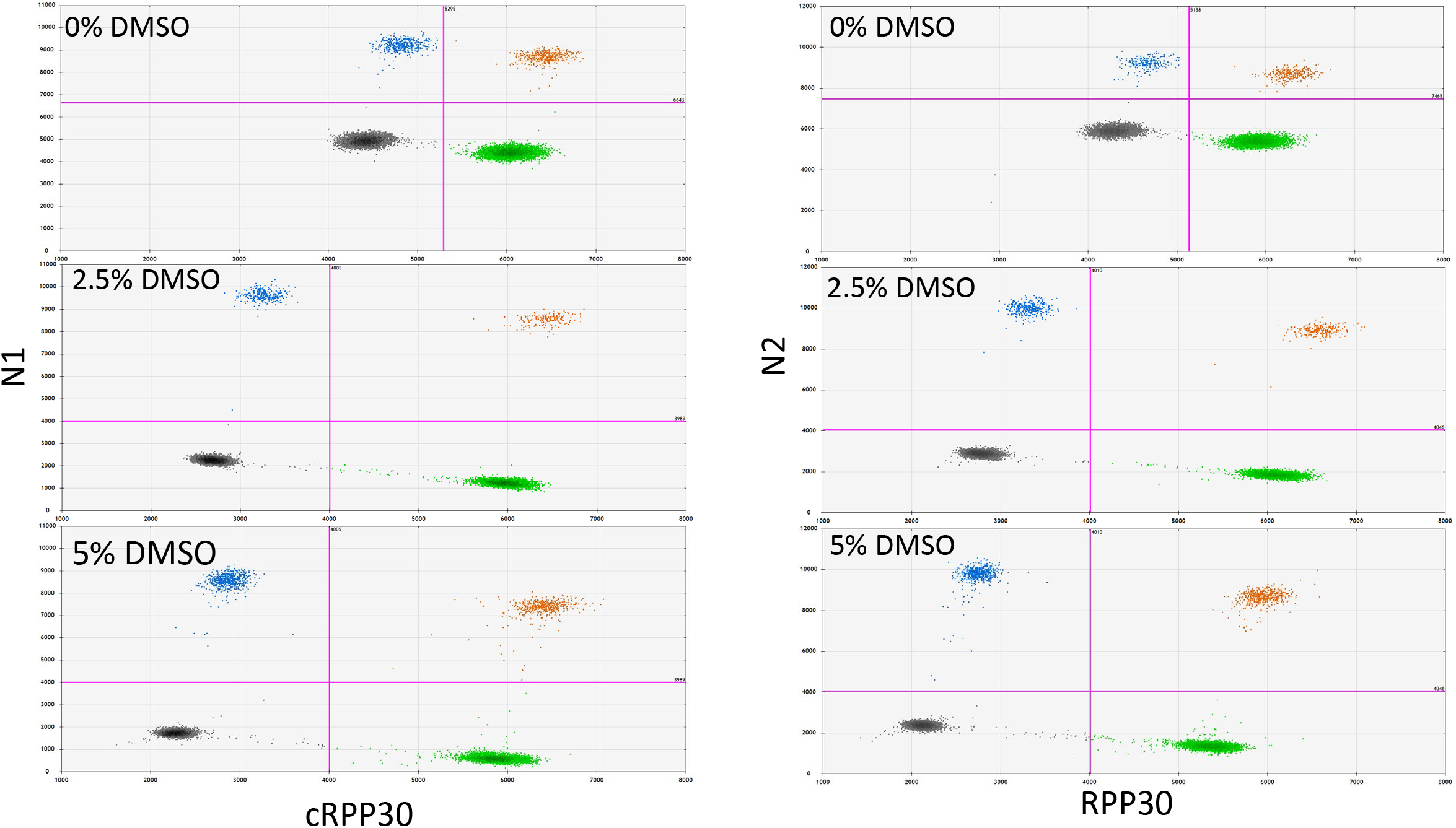
DMSO decreases negative droplet intensity in RT-ddPCR assays. The RT-ddPCR reactions containing 0%, 2.5%, or 5% DMSO were performed for a Chelex-lowTE sample prepared with the ATCC SARS-CoV-2 virions and 293FT cells using N1 and cRPP30 (left panel) or N2 and RPP30 (right panel). The grey clusters represent negative droplets; blue and green clusters represent Fam and Hex positive droplets, respectively; and orange clusters represent double positive droplets.

**Figure S2.**
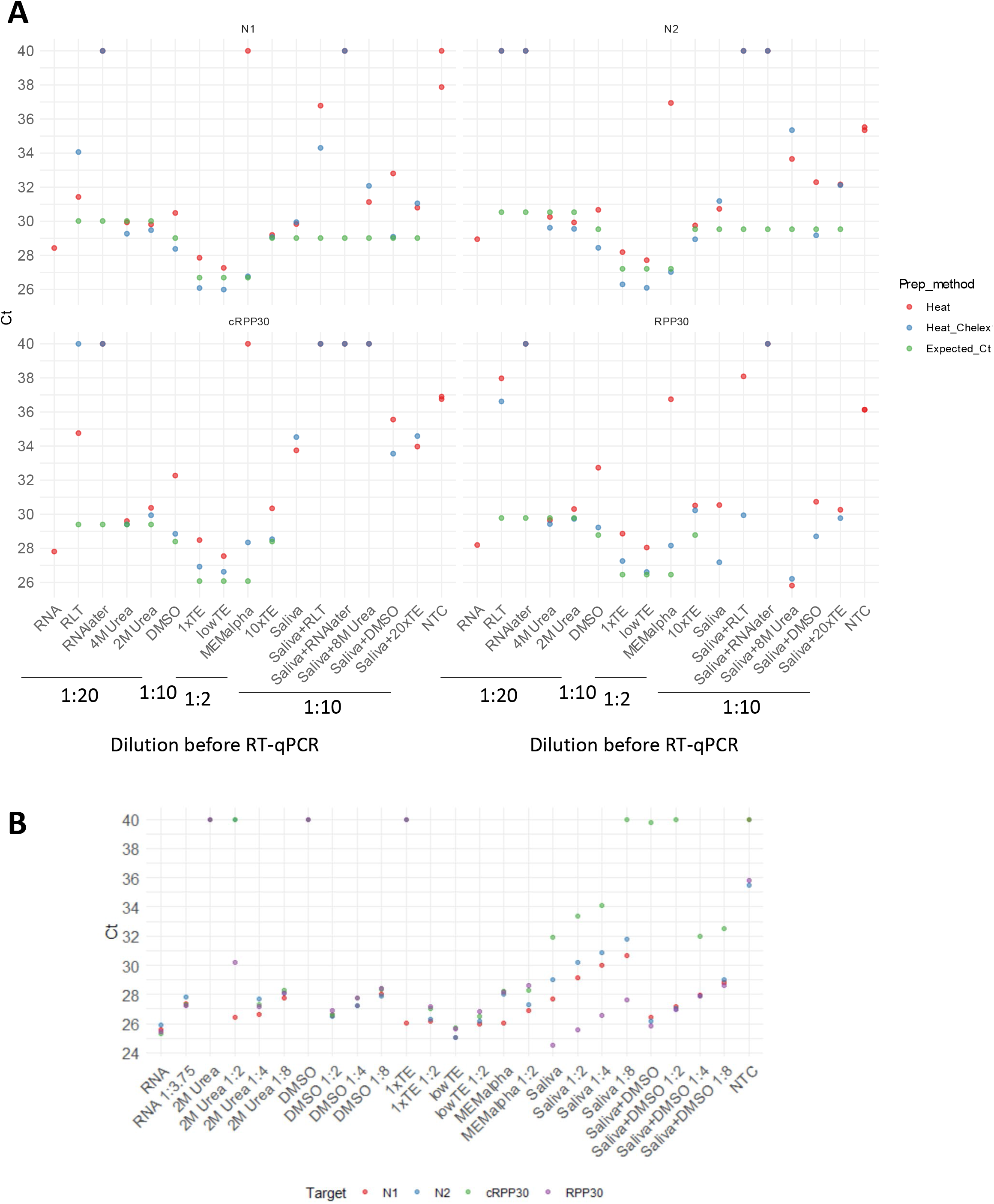
SARS-CoV-2 prepared in different buffers used for RT-qPCR without RNA-extraction. **(A)** Samples were diluted in H2O as indicated at the bottom. Expected_Ct refers to Ct calculated based on Ct from extracted RNA normalized with added virion numbers after dilution using the ΔCt method. **(B)** Buffer compatibility in RT-qPCR. Sample RNA, not heated and 5 µl of which contained materials extracted from 6,250 virions. Other samples were heated in the presence of Chelex, of which undiluted samples also contained 6,250 virions per 5 µl. Samples were diluted in H2O. Samples with undetermined Ct values were plotted as Ct 40. The NEB Luna RT-qPCR kit and NEB-Luna-Program I was used. NTC, no-template control.

**Figure S3.**
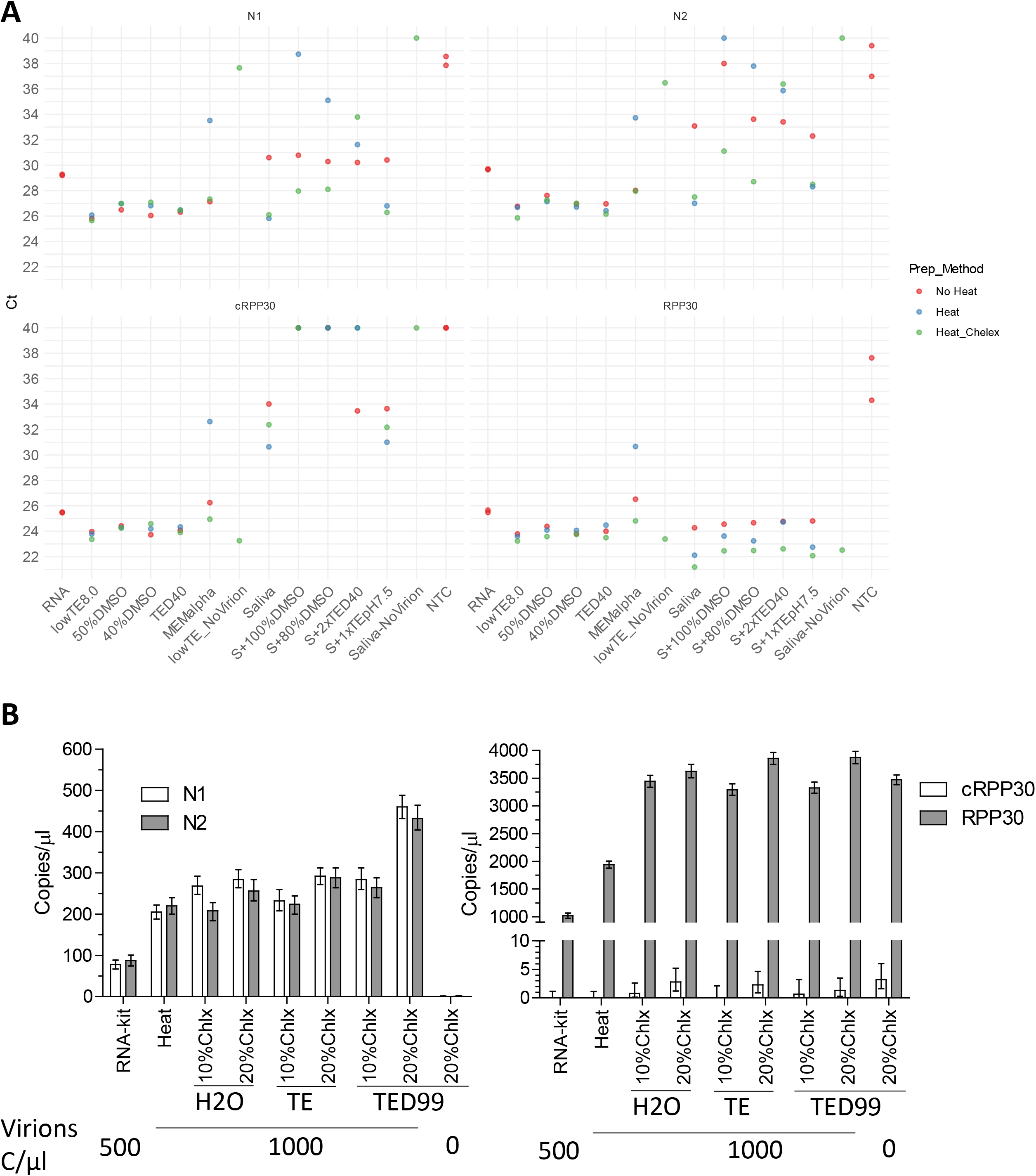
Tris EDTA and DMSO containing buffers. **(A)** RT-qPCR of samples with heat-inactivated ATCC SARS-CoV-2 virions. 5 µl of samples were used for one reaction in RT-qPCR except that samples in MEM α were diluted 1:1 with H2O. Samples with undetermined Ct values were plotted as Ct 40. The NEB Luna RT-qPCR kit and NEB-Luna-Program I was used. **(B)** RT-ddPCR of saliva samples with heat-inactivated ATCC SARS-CoV-2 virions. The Chelex was prepared in H2O, lowTE or TED99 (lowTE with 99% DMSO). RNA-kit refers to RNA extracted with the RNeasy Protect Saliva Mini Kit. NTC, no-template control.

**Figure S4.**
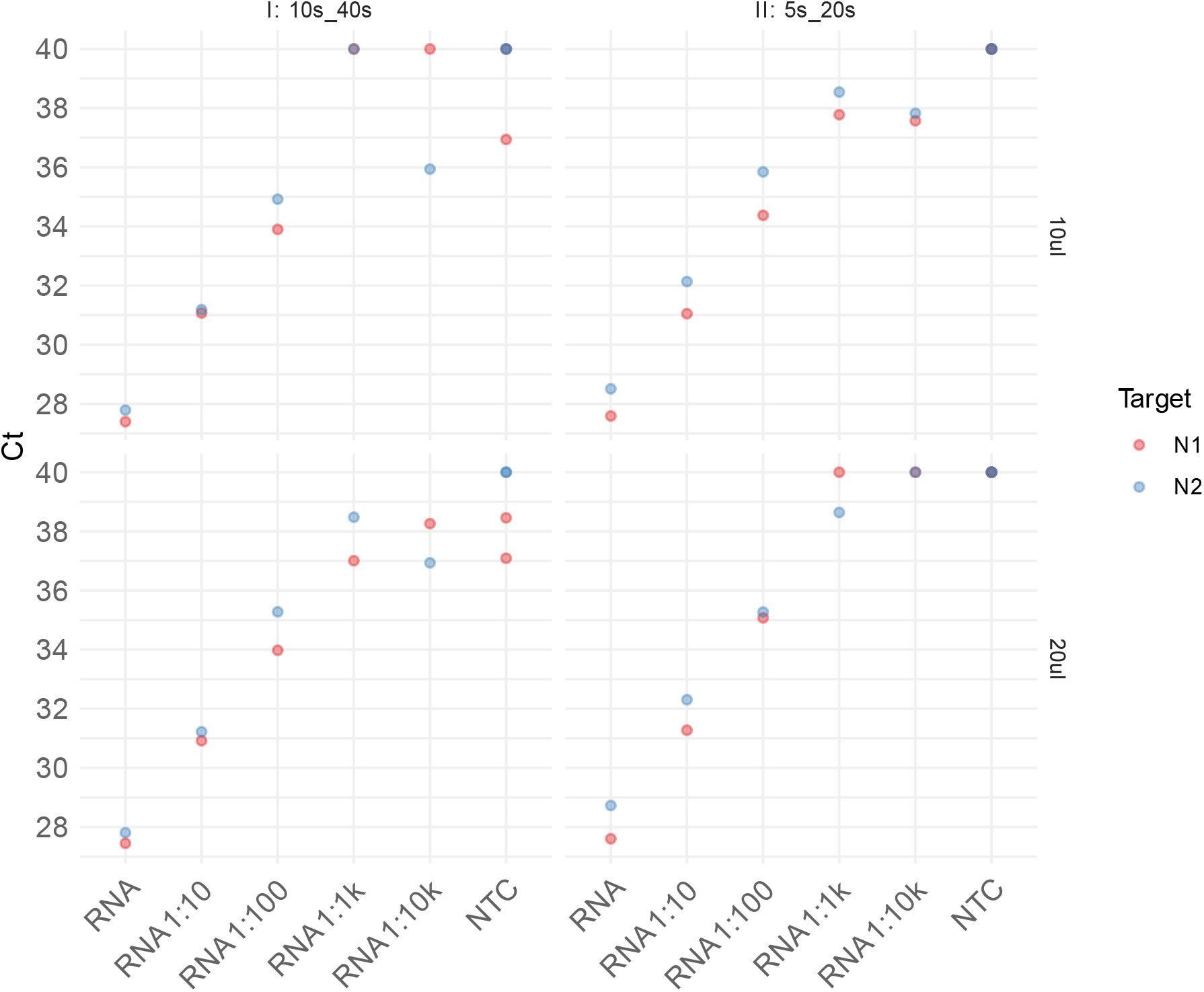
Optimization of the NEB Luna RT-qPCR assay. Extracted RNA samples were serial diluted and assayed either using 2.5 µl sample in a 10 reaction volume or 5 µl in a 20 µl reaction, and using a longer PCR protocol (I: 10 seconds of denature and 40 seconds of annealing/extension) or a shorter PCR protocol (II: 5 seconds of denature and 20 seconds of annealing/extension). NTC, no-template control.

